# Alcohol Use and Sustained Virologic Response to Hepatitis C Virus Direct-Acting Antiviral Therapy: A National Observational Cohort Study

**DOI:** 10.1101/2022.11.06.22281998

**Authors:** Emily J. Cartwright, Chloe Pierret, Caroline Minassian, Denise A. Esserman, Janet P. Tate, Matthew B. Goetz, Debika Bhattacharya, David A. Fiellin, Amy C. Justice, Vincent Lo Re, Christopher T. Rentsch

**Affiliations:** Division of Infectious Diseases, Emory University School of Medicine, Atlanta, GA, US; Atlanta Veterans Affairs Medical Center, Decatur, GA, US; Faculty of Epidemiology and Population Health, London School of Hygiene & Tropical Medicine, London, UK; Veterans Affairs Connecticut Healthcare System, US Department of Veterans Affairs, West Haven, CT, US; Department of Internal Medicine, Yale School of Medicine, New Haven, CT, US; Department of Internal Medicine, David Geffen School of Medicine, University of California, Los Angeles, Los Angeles, CA, US; Veterans Affairs Greater Los Angeles Health Care System, US Department of Veterans Affairs, Los Angeles, CA, US; Yale Program in Addiction Medicine, Yale School of Medicine, New Haven, CT, US; Yale School of Public Health, New Haven, CT, US; Division of Infectious Diseases, Department of Medicine and Center for Clinical Epidemiology and Biostatistics, Perelman School of Medicine, University of Pennsylvania, Philadelphia, PA, US

## Abstract

**Background:** Some payors and clinicians require alcohol abstinence for direct-acting antiviral (DAA) therapy for chronic hepatitis C virus (HCV) infection.

**Objective:** To evaluate whether alcohol use at DAA treatment initiation was associated with decreased odds of sustained virologic response (SVR).

**Design:** Observational cohort study using electronic health records.

**Setting:** US Department of Veterans Affairs (VA), the largest integrated national healthcare system that provides unrestricted access to HCV treatment.

**Patients:** All patients born between 1945 and 1965 who were dispensed DAA therapy between 1 January 2014 and 30 June 2018.

**Measurements:** We used multivariable logistic regression to estimate odds ratios (ORs) and 95% confidence intervals (CIs) of SVR associated with alcohol category. SVR was defined as undetectable HCV RNA ≥12 weeks after completion of DAA therapy. Alcohol category was determined using information on alcohol use disorder diagnoses and Alcohol Use Disorders Identification Test - C (AUDIT-C) at DAA initiation.

**Results:** Among 69,229 patients who initiated DAA therapy (mean age 63 years; 97% men; 50% non-Hispanic White; 41% non-Hispanic Black; 85% HCV genotype 1), 65,355 (94.4%) of patients achieved SVR. After multivariable adjustment, we found no difference in SVR across alcohol use categories (lowest OR 0.92, 95% CI 0.82-1.04). There was no evidence of interaction by stage of hepatic fibrosis measured by FIB-4 (p-interaction=0.3001).

**Limitations:** Predominately male population.

**Conclusion:** Alcohol use was not associated with lower odds of SVR, suggesting that DAA therapy should not be withheld due to alcohol use. Restricting access to DAA therapy based on alcohol use creates an unnecessary barrier to patients and challenges HCV elimination goals.

**Funding source:** National Institute on Alcohol Abuse and Alcoholism

## Introduction

Previously, chronic hepatitis C virus (HCV) infection was treated with interferon-based regimens; a poorly tolerated therapy. In the interferon era, persons with active alcohol use in the previous year were more likely to discontinue interferon-based HCV treatment, and many clinicians were reluctant to treat patients with recent alcohol use.^1^ However, for those persons who successfully completed interferon-based therapy, comparable rates of sustained virologic response (SVR) were achieved regardless of reported alcohol use.^1–4^ In 2009, the American Association for the Study of Liver Diseases treatment guidelines stated that candidates for HCV treatment should be abstinent from alcohol for a minimum period of 6 months prior to initiating treatment.^5^ The Department of Veterans Affairs HCV treatment recommendations required referral to an addiction specialist prior to treatment initiation but did not recommend withholding HCV antiviral therapy on the basis of alcohol consumption.^6^ With the advent of safe and highly effective direct-acting antiviral (DAA) therapy for HCV, the impact of alcohol use on achieving SVR is less clear.

Tsui and colleagues published an analysis examining the relationship between alcohol use and SVR in a cohort of US Veterans who initiated DAA therapy between 2014 and mid-2015.^7^ The study found relatively high rates of SVR (>91%) across all alcohol use categories and no association between alcohol use and SVR in all primary analyses. However, this study included patients who reported abstinence from alcohol as the referent group, which is a heterogeneous group known to include persons who consumed alcohol previously and quit due to alcohol-related or other health problems (“sick quitters”).^8^ Analyses that combine prior drinkers who are abstinent and lifetime alcohol abstainers in the same referent group may be prone to biased results.

Current American Association for the Study of Liver Diseases/Infectious Diseases Society of America guidelines for the treatment of chronic HCV infection advise that patients with HCV avoid excess alcohol use but do not recommend restricting access to DAA therapy based on alcohol intake regardless of any level of consumption. Similarly, the Department of Veterans Affairs (VA) – the largest provider of HCV care in the United States – does not recommend withholding DAA therapy from patients with alcohol use disorder and DAA therapy is available to patients with HCV infection at no or substantially reduced cost to the individual in the VA system.^9,10^ Despite these recommendations, some clinicians continue to delay or withhold HCV therapy from patients with ongoing alcohol use.^11–13^. Furthermore, some payors include alcohol abstinence as a requirement for reimbursement of DAA therapy for HCV.^14^ We used national VA electronic health record data to examine the relationship between alcohol consumption and SVR by distinguishing individuals with a history of alcohol use disorder among those who report current alcohol abstinence.

## Methods

### Study design and data source

We conducted a retrospective cohort study using electronic health record data from the VA 1945-1965 Birth Cohort, which includes all individuals born between 1945 and 1965 who had at least one VA encounter on or after 1 October 1999. The VA comprises more than 1200 points of healthcare nationwide, including hospitals, medical centers, and community outpatient clinics, from which all care is recorded in a central data repository, with daily uploads into the Veterans Affairs Corporate Data Warehouse. We chose to use data from the VA 1945-1965 Birth Cohort because persons born between 1945 and 1965 have a 6-fold higher prevalence of HCV infection compared to all other age groups, estimated at 7.5% in 2016.^15^ The VA 1945-1965 Birth Cohort includes data on demographics, outpatient and inpatient encounters, International Classification of Disease 9^th^ and 10^th^ edition (ICD-9 and ICD-10) diagnostic codes, smoking and alcohol use, pharmacy dispensing records, laboratory measures, vital signs, and death.

This study is reported as per the Strengthening the Reporting of Observational Studies in Epidemiology (STROBE) and REporting of studies Conducted using Observational Routinely-collected health Data (RECORD) statements (see **Appendix**).

### Study population

We included all patients who initiated interferon-free DAA therapy between 1 January 2014 and 30 June 2018. The index date was defined as the day the patient was dispensed their first DAA regimen, including daclatasvir, dasabuvir, elbasvir, glecaprevir, grazoprevir, ledipasvir, ombitasvir, paritaprevir, pibrentasvir, simeprevir, sofosbuvir, velpatasvir, or voxileprevir. Ribavirin and ritonavir were also considered part of a DAA regimen when they were prescribed concurrently with sofosbuvir, paritaprevir/ombitasvir. We have previously demonstrated that DAA therapies are accurately recorded in VA electronic health record data (positive predictive value 98.6% and negative predictive value 99.0%).^16^

### Alcohol category

Our primary exposure variable combined information on alcohol consumption and alcohol use disorder (AUD) ascertained in the 18 months prior to the index date. Alcohol consumption was assessed using the Alcohol Use Disorders Identification Test – Consumption (AUDIT-C), a three-item questionnaire that ascertains quantity and frequency of alcohol use to detect heavy drinking and/or active alcohol use disorder.^17,18^ Since 2007, the VA has required annual AUDIT-C screening on all patients during routine healthcare visits in primary care.^19^ AUDIT-C scores range from 0 to 12 with the likelihood of physiologic injury and mortality increasing with higher AUDIT-C score.^20^ We classified patients as having AUD by the presence of at least one inpatient or outpatient diagnostic code, including ICD-9/-10 codes: 303.*, 305.0*, F10.1* excluding F10.13*, or F10.2* excluding F10.21. Both AUDIT-C and AUD were ascertained prior to index date. We used the AUDIT-C measure closest to the index date for patients with more than one available measurement in the 18-month ascertainment window.

We classified patients into five exhaustive and mutually exclusive groups based on their proximal AUDIT-C measure and presence/absence of AUD: 1) abstinent with no AUD (AUDIT-C=0 and absence of AUD diagnosis); 2) abstinent with AUD (AUDIT-C=0 and presence of AUD diagnosis); 3) lower-risk consumption (AUDIT-C 1-3 and absence of AUD diagnosis); 4) moderate-risk consumption (AUDIT-C 4-7 and absence of AUD diagnosis); and 5) high-risk consumption or AUD (AUDIT-C ≥8 or presence of AUD diagnosis with non-zero AUDIT-C). We used lower-risk consumption as the referent group in all analyses since patients who report no current alcohol use (AUDIT-C=0) in the VA are a heterogeneous group comprising very few lifetime abstainers and the majority of whom quit drinking after alcohol-related or health problem.^8^ Thus, using abstinent individuals as the referent group increases the risk of misclassification, confounding, and weaker associations, especially among middle-aged and older adults.

### Outcome

The primary outcome was SVR defined by undetectable HCV RNA ≥12 weeks after completion of DAA therapy. To mitigate the potential for capturing re-infections, we only considered HCV RNA measurements up to 6 months after completion of DAA therapy to define SVR. Therefore, HCV RNA measurements through 31 December 2018 were extracted for analysis. If there were multiple HCV RNA measurements available, the latest result was chosen. For patients with no measurements ≥12 weeks after completion of DAA therapy, we selected the latest result available in the 4 to 11 weeks after completion of DAA therapy. While the gold standard for HCV cure has historically been SVR at ≥24 weeks,^21^ previous studies have demonstrated high concordance between SVR defined at 4-, 12- and 24-weeks post treatment.^22^

### Covariates

**Figure 1** depicts the study design and details on exposure and covariate ascertainment windows. Covariates included age, sex, race/ethnicity, rural/urban residence type, body mass index, smoking status, HIV co-infection, Charlson Comorbidity Index, liver-related variables (i.e., HCV genotype, hepatitis B co-infection, fibrosis 4 [FIB-4] score, hepatic decompensation, liver cancer), and other HCV treatment-related variables (i.e., previous receipt of non-DAA therapy for HCV infection, year of DAA therapy initiation, DAA regimen type). Rural/urban residence type was defined using geographic information system coding based upon established criteria.^23^ Body mass index was calculated using the most recent height and weight measures in the three years before index date. Smoking status was determined by the most frequent response in the five years before index date. Presence of clinical comorbidities (i.e., Charlson Comorbidity Index, hepatic decompensation, liver cancer) was determined by one inpatient or two outpatient diagnoses using ICD-9 or ICD-10 codes in the five years before index date, except for HIV co-infection, which was considered present if diagnosed ever before index date. Hepatitis B was defined using either ICD-9/-10 codes or a positive surface antigen test in the five years before index date. FIB-4 scores were calculated using the most recent alanine aminotransferase, aspartate aminotransferase, and platelet count measures in the 18 months before index date using a validated algorithm.^24^

**Figure 1.**
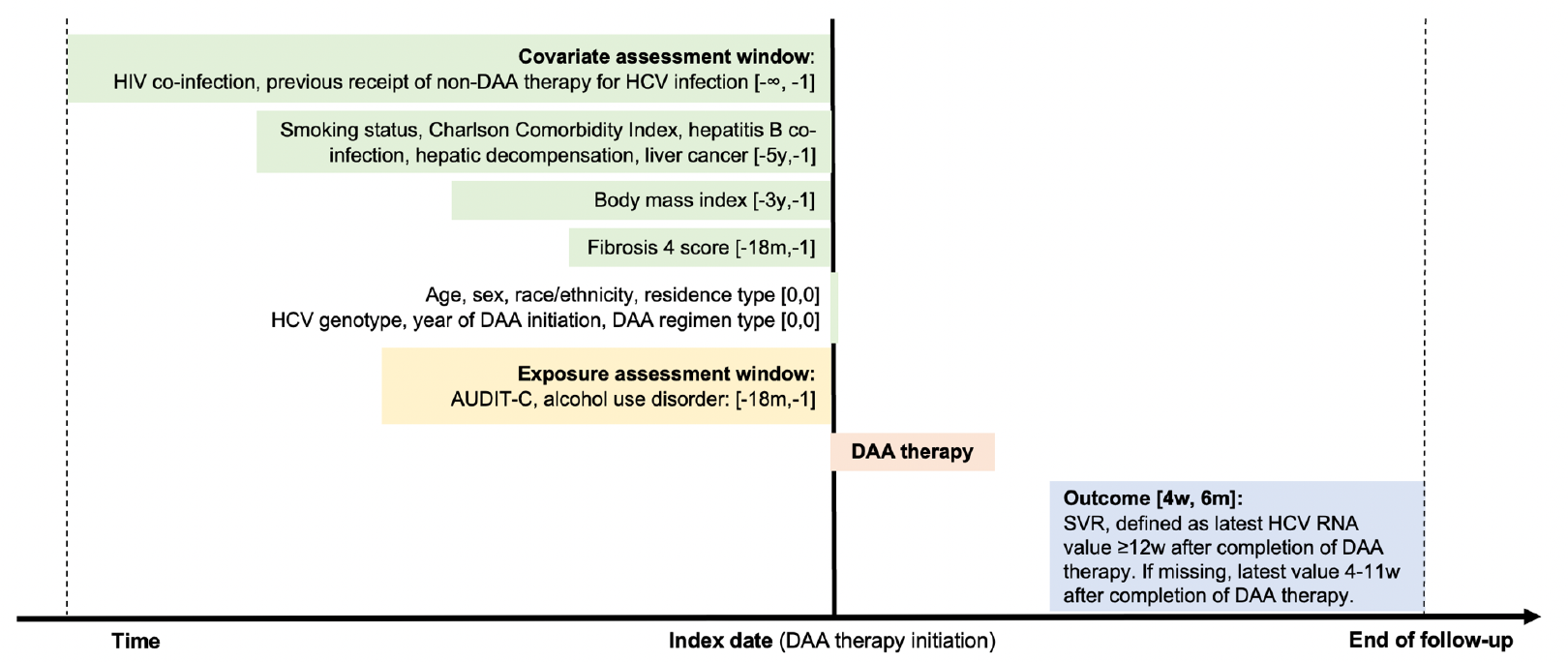
Study diagram *Abbreviations:* HIV. human immunodeficiency virus; DAA, direct-acting antiviral; HCV, hepatitis C virus; AUDIT-C, Alcohol Use Disorder Identification Test-Consumption; SVR. sustained virologic response: y, year; m, month; w, week

### Statistical analysis

We calculated absolute standardized mean differences (SMDs) to identify differences in the covariate distribution between patients with and without AUDIT-C or HCV RNA labs to define SVR. We considered SMD<0.2 as balanced;^25^ thus, SMD≥0.2 suggested meaningful imbalance in characteristics.

After excluding those with missing alcohol and HCV RNA results, an additional 7,339 (9.6%) had missing data for any of the other covariates (i.e., 6.9% missing FIB-4, 1.4% missing HCV genotype, 1.1% missing smoking status, and 0.8% missing body mass index). Our primary analyses were performed on complete cases because the overall level of missingness among covariates was <10% and a large proportion of missingness was likely to be missing not at random. In this circumstance, although multiple imputation is not appropriate, a complete case analysis will be unbiased if, conditional on model covariates, missingness is independent of the outcome.^26^

We characterized the cohort of complete cases by displaying the distribution of covariates by alcohol category. We then used logistic regression to estimate odds ratios (ORs) and 95% confidence intervals (CIs) for the association between alcohol category and SVR. First, we estimated crude associations in a model including all patients with exposure and outcome data. Second, we estimated crude associations in a model including complete cases. Third, we fitted a multivariable model fully adjusted for demographics, clinical characteristics, liver-related variables; and other HCV treatment-related variables. FIB-4 scores >3.25 predict advanced liver fibrosis/cirrhosis and risk of liver cancer;^27,28^ therefore, we assessed the possible interaction between alcohol category and FIB-4 score in the fully adjusted model.

### Sensitivity analyses

We compared estimates from the fully adjusted model in the primary analysis to models including patients with missing outcome data under three separate assumptions: 1) multiple imputation (ten imputations) of the outcome with the imputation model including all extracted covariates; 2) all patients with missing outcome data were assumed to have achieved SVR; and 3) all patients with missing outcome data were assumed to not have achieved SVR. Although multiple imputation assumes data to define SVR were missing at random,^29^ we included this sensitivity analysis to compare our findings with previous evidence.

## Results

### Cohort description

Among 94,388 patients who initiated DAA therapy between 1 January 2014 and 30 June 2018, 3,763 (4.0%) had missing AUDIT-C and 14,057 (15.5%) had missing HCV RNA labs to define SVR within 4 weeks to 6 months after DAA completion. There was a small difference in Charlson Comorbidity Index scores between patients with and without AUDIT-C (SMD=0.29); however, there was little to no difference between patients with and without AUDIT-C (**eTable 1**) or those with and without HCV RNA labs (**eTable 2**) in the distribution for all other covariates (all SMD<0.2 with majority SMD<0.1).

Of the 69,229 patients who were included in the primary analyses, median age was 62.8 years (interquartile range 59.4-66.0 years), 67,150 (97.0%) were men, 34,655 (50.1%) were non-Hispanic White, 28,094 (40.6%) were non-Hispanic Black, and 46,220 (66.8%) were current smokers at DAA initiation (**Table 1**). Most patients had HCV genotype 1 (n=58,477; 84.5%), some had previous receipt of non-DAA therapy for HCV infection (n=10,632; 15.4%), and few had HIV co-infection (n=2,217; 3.2%) or HBV co-infection (n=1,308; 1.9%). About half of patients (n=35,032; 50.6%) had FIB-4 scores between 1.45 and 3.25, while 16,103 (23.3%) patients had scores below 1.45 and 18,094 (26.1%) patients had scores above 3.25 at DAA initiation. Sofosbuvir/ledipasvir (58.8%) was the most frequently prescribed DAA regimen.

**Table 1.** Characteristics of 69,229 patients who initiated direct-acting antiviral therapy by alcohol category

|  | Abstinent with<br>no AUD<br>n=32,290 | Abstinent with<br>AUD<br>n=9,192 | Lower-risk<br>consumption<br>n=13,415 | Moderate-risk<br>consumption<br>n=3,117 | High-risk<br>consumption<br>or AUD<br>n=11,215 |
| --- | --- | --- | --- | --- | --- |
| **Demographics** |  |  |  |  |  |
| Age in years, median (IQR) | 63.3 (59.9-66.4) | 61.6 (58.4-65.0) | 63.0 (59.7-66.1) | 63.0 (59.8-66.1) | 61.8 (58.6-65.2) |
| 48-54 | 1,440 (4.5) | 725 (7.9) | 662 (4.9) | 143 (4.6) | 793 (7.1) |
| 55-59 | 6,787 (21.0) | 2,635 (28.7) | 2,982 (22.2) | 688 (22.1) | 3,212 (28.6) |
| 60-64 | 12,418 (38.5) | 3,497 (38.0) | 5,233 (39.0) | 1,248 (40.0) | 4,290 (38.3) |
| 65-73 | 11,645 (36.1) | 2,335 (25.4) | 4,538 (33.8) | 1,038 (33.3) | 2,920 (26.0) |
| Sex |  |  |  |  |  |
| Women | 1,074 (3.3) | 251 (2.7) | 474 (3.5) | 49 (1.6) | 231 (2.1) |
| Men | 31,216 (96.7) | 8,941 (97.3) | 12,941 (96.5) | 3,068 (98.4) | 10,984 (97.9) |
| Race/ethnicity |  |  |  |  |  |
| Non-Hispanic White | 16,403 (50.8) | 4,666 (50.8) | 6,536 (48.7) | 1,723 (55.3) | 5,327 (47.5) |
| Non-Hispanic Black | 12,702 (39.3) | 3,738 (40.7) | 5,608 (41.8) | 1,117 (35.8) | 4,929 (44.0) |
| Hispanic | 1,649 (5.1) | 450 (4.9) | 638 (4.8) | 112 (3.6) | 506 (4.5) |
| Other/missing | 1,536 (4.8) | 338 (3.7) | 633 (4.7) | 165 (5.3) | 453 (4.0) |
| Residence type |  |  |  |  |  |
| Rural | 8,815 (27.3) | 2,099 (22.8) | 3,516 (26.2) | 964 (30.9) | 2,695 (24.0) |
| Urban | 23,475 (72.7) | 7,093 (77.2) | 9,899 (73.8) | 2,153 (69.1) | 8,520 (76.0) |
| **Clinical characteristics** |  |  |  |  |  |
| Body mass index |  |  |  |  |  |
| Underweight | 467 (1.4) | 126 (1.4) | 208 (1.6) | 58 (1.9) | 243 (2.2) |
| Normal | 8,482 (26.3) | 2,601 (28.3) | 3,783 (28.2) | 1,084 (34.8) | 3,952 (35.2) |
| Overweight | 12,244 (37.9) | 3,515 (38.2) | 5,232 (39.0) | 1,186 (38.0) | 4,208 (37.5) |
| Obese | 11,097 (34.4) | 2,950 (32.1) | 4,192 (31.2) | 789 (25.3) | 2,812 (25.1) |
| Smoking status |  |  |  |  |  |
| Never | 5,117 (15.8) | 795 (8.6) | 1,840 (13.7) | 323 (10.4) | 779 (6.9) |
| Current | 18,686 (57.9) | 7,053 (76.7) | 8,831 (65.8) | 2,305 (73.9) | 9,345 (83.3) |
| Former | 8,487 (26.3) | 1,344 (14.6) | 2,744 (20.5) | 489 (15.7) | 1,091 (9.7) |
| HIV co-infection | 1,072 (3.3) | 318 (3.5) | 409 (3.0) | 74 (2.4) | 344 (3.1) |
| Charlson Comorbidity Index |  |  |  |  |  |
| 1 | 12,107 (37.5) | 3,322 (36.1) | 6,031 (45.0) | 1,578 (50.6) | 4,883 (43.5) |
| 2 | 8,230 (25.5) | 2,394 (26.0) | 3,398 (25.3) | 834 (26.8) | 2,976 (26.5) |
| 3 | 3,273 (10.1) | 879 (9.6) | 1,284 (9.6) | 265 (8.5) | 1,072 (9.6) |
| 4 | 3,313 (10.3) | 913 (9.9) | 1,098 (8.2) | 195 (6.3) | 921 (8.2) |
| ≥5 | 5,367 (16.6) | 1,684 (18.3) | 1,604 (12.0) | 245 (7.9) | 1,363 (12.2) |
| **Liver-related variables** |  |  |  |  |  |
| HCV Genotype |  |  |  |  |  |
| 1 | 27,342 (84.7) | 7,727 (84.1) | 11,358 (84.7) | 2,584 (82.9) | 9,466 (84.4) |
| 2 | 2,889 (8.9) | 824 (9.0) | 1,242 (9.3) | 354 (11.4) | 1,023 (9.1) |
| 3 | 1,739 (5.4) | 560 (6.1) | 663 (4.9) | 153 (4.9) | 610 (5.4) |
| 4/5/6 | 320 (1.0) | 81 (0.9) | 152 (1.1) | 26 (0.8) | 116 (1.0) |
| Hepatitis B co-infection | 581 (1.8) | 253 (2.8) | 185 (1.4) | 43 (1.4) | 246 (2.2) |
| Fibrosis 4 score |  |  |  |  |  |
| <1.45 | 7,402 (22.9) | 2,162 (23.5) | 3,400 (25.3) | 712 (22.8) | 2,427 (21.6) |
| 1.45-3.25 | 16,489 (51.1) | 4,353 (47.4) | 7,137 (53.2) | 1,641 (52.6) | 5,412 (48.3) |
| >3.25 | 8,399 (26.0) | 2,677 (29.1) | 2,878 (21.5) | 764 (24.5) | 3,376 (30.1) |
| Hepatic decompensation | 1,091 (3.4) | 640 (7.0) | 117 (0.9) | 14 (0.4) | 346 (3.1) |
| Liver cancer | 786 (2.4) | 363 (3.9) | 154 (1.1) | 34 (1.1) | 200 (1.8) |
| **Treatment-related variables** |  |  |  |  |  |
| Previous non-DAA therapy for HCV infection | 6,021 (18.6) | 1,551 (16.9) | 1,635 (12.2) | 298 (9.6) | 1,127 (10.0) |
| Year of DAA therapy initiation |  |  |  |  |  |
| 2014 | 3,298 (10.2) | 978 (10.6) | 760 (5.7) | 116 (3.7) | 466 (4.2) |
| 2015 | 9,995 (31.0) | 3,111 (33.8) | 3,556 (26.5) | 584 (18.7) | 2,623 (23.4) |
| 2016 | 11,574 (35.8) | 3,325 (36.2) | 5,204 (38.8) | 1,271 (40.8) | 4,502 (40.1) |
| 2017 | 5,932 (18.4) | 1,400 (15.2) | 3,099 (23.1) | 923 (29.6) | 2,826 (25.2) |
| 2018 | 1,491 (4.6) | 378 (4.1) | 796 (5.9) | 223 (7.2) | 798 (7.1) |
| DAA regimen type |  |  |  |  |  |
| Sofosbuvir/ledipasvir | 18,526 (57.4) | 5,569 (60.6) | 7,954 (59.3) | 1,867 (59.9) | 6,805 (60.7) |
| Elbasvir/grazoprevir | 3,387 (10.5) | 729 (7.9) | 1,484 (11.1) | 361 (11.6) | 1,234 (11.0) |
| Sofosbuvir/velpatasvir ± voxileprevir | 2,270 (7.0) | 592 (6.4) | 1,085 (8.1) | 341 (10.9) | 1,000 (8.9) |
| Glecaprevir/pibrentasvir | 972 (3.0) | 234 (2.5) | 552 (4.1) | 127 (4.1) | 506 (4.5) |
| Paritaprevir/ritonavir/ombitasvir ± dasabuvir | 2,747 (8.5) | 697 (7.6) | 1,084 (8.1) | 199 (6.4) | 725 (6.5) |
| Sofosbuvir/daclatasvir | 389 (1.2) | 121 (1.3) | 148 (1.1) | 28 (0.9) | 124 (1.1) |
| Simeprevir/sofosbuvir | 1,410 (4.4) | 427 (4.6) | 295 (2.2) | 29 (0.9) | 205 (1.8) |
| Sofosbuvir/ribavirin | 2,589 (8.0) | 823 (9.0) | 813 (6.1) | 165 (5.3) | 616 (5.5) |

*Abbreviations:* AUD, alcohol use disorder; IQR, interquartile range; HIV, human immunodeficiency virus; HCV, hepatitis C virus; DAA, direct-acting antiviral
*Notes:* All statistics reported as n(%) unless otherwise noted.

### Alcohol category and SVR

According to the five alcohol categories, patients were: 32,290 (46.6%) abstinent with no AUD, 9,192 (13.3%) abstinent with AUD, 13,415 (19.4%) lower-risk consumption, 3,117 (4.5%) moderate-risk consumption, and 11,215 (16.2%) high-risk consumption or AUD. Overall, 65,355 (94.4%) of all DAA-initiating patients achieved SVR, with 58,651 SVR outcomes measured 12 weeks to 6 months after DAA completion, and 6,704 SVR outcomes measured in the 4 to 12 weeks after DAA completion.

In an unadjusted model, patients who were abstinent with no AUD history (OR 0.89, 95% CI 0.81-0.97) and those who were abstinent with AUD history (OR 0.71, 95% CI 0.64-0.80) at DAA initiation had lower odds of achieving SVR compared with patients who reported lower-risk consumption (**Table 2**). However, after adjusting for demographics, clinical characteristics, liver-related variables, and HCV treatment-related variables, we found no evidence that any alcohol category was significantly associated with decreased odds of SVR (OR 1.09, 95% CI 0.99-1.20 for abstinent with no AUD history; OR 0.92, 95% CI 0.82-1.04 for abstinent with AUD history; OR 0.96, 95% CI 0.80-1.15 for moderate-risk consumption; OR 0.95, 95% CI 0.85-1.07 for high-risk consumption or AUD) compared with patients who reported lower-risk consumption (**Table 2**). In addition, we found no evidence that the association between alcohol category and odds of SVR differed by baseline stage of hepatic fibrosis measured by FIB-4 less than 3.25 versus greater than 3.25 (p-interaction=0.3001; **Table 2**).

**Table 2.** Associations between alcohol category and sustained virologic response

|  | All patients with<br>exposure and outcome<br>data (n=76,568) | Complete case analysis<br>(n=69,229) |  | By FIB-4 score (p-interaction=0.3001) |  |
| --- | --- | --- | --- | --- | --- |
|  | Unadjusted | Unadjusted | Fully adjusted | ≤3.25 (n=51,135) | >3.25 (n=18,094) |
| Abstinent with no AUD | 0.88 (0.80-0.95) | 0.89 (0.81-0.97) | 1.09 (0.99-1.20) | 1.11 (0.99-1.25) | 1.08 (0.92-1.27) |
| Abstinent with AUD | 0.70 (0.63-0.78) | 0.71 (0.64-0.80) | 0.92 (0.82-1.04) | 0.96 (0.82-1.11) | 0.88 (0.72-1.06) |
| Lower-risk consumption | ref | ref | ref | ref | ref |
| Moderate-risk consumption | 1.03 (0.87-1.22) | 1.04 (0.86-1.24) | 0.96 (0.80-1.15) | 1.00 (0.79-1.26) | 0.87 (0.64-1.19) |
| High-risk consumption or AUD | 0.91 (0.81-1.01) | 0.92 (0.82-1.03) | 0.95 (0.85-1.07) | 0.91 (0.79-1.06) | 1.02 (0.82-1.21) |

*Abbreviations:* AUD, alcohol use disorder
*Notes:* Fully adjusted model included age, sex, race/ethnicity, residence type, body mass index, smoking status, HIV co-infection, Charlson Comorbidity Index, liver-related variables (i.e., HCV genotype, hepatitis B co-infection, fibrosis 4 score, hepatic decompensation, liver cancer, and other HCV treatment-related variables (i.e., previous receipt of non-DAA therapy for HCV infection, year of DAA therapy initiation, DAA regimen type).

### Sensitivity analyses

Findings remained the same after including patients with missing data using multiple imputation (lowest OR 0.90, 95% CI 0.80-1.02 for patients who were abstinent with AUD history) and assuming all patients with missing outcome data achieved SVR (lowest OR 0.94, 95% CI 0.84-1.06 for patients who were abstinent with AUD history; **Table 3**). In an analysis assuming the highly unlikely scenario that all patients with missing outcome data did not achieve SVR, we observed patients who were abstinent with AUD history (OR 0.92, 95% CI 0.86-0.98) and those with high-risk consumption or AUD (OR 0.88, 95% CI 0.83-0.93) had lower odds of SVR compared to patients who reported lower-risk consumption. However, the confidence limits for these associations were close to the null.

**Table 3.** Sensitivity analyses using various assumptions about patients with missing outcome data

|  | Complete case analysis<br>(n=69,229) | Sensitivity analyses (n=81,703) |  |  |
| --- | --- | --- | --- | --- |
|  | Fully adjusted | Multiple imputation<br>of missing SVR | Missing SVR<br>assumed to be cured | Missing SVR<br>assumed to be not<br>cured |
| Abstinent with no AUD | 1.09 (0.99-1.20) | 1.08 (0.97-1.20) | 1.09 (0.99-1.20) | 1.00 (0.96-1.05) |
| Abstinent with AUD | 0.92 (0.82-1.04) | 0.90 (0.80-1.02) | 0.94 (0.84-1.06) | 0.92 (0.86-0.98) |
| Lower-risk consumption | ref | ref | ref | ref |
| Moderate-risk consumption | 0.96 (0.80-1.15) | 0.95 (0.76-1.19) | 0.95 (0.79-1.15) | 1.03 (0.94-1.13) |
| High-risk consumption or AUD | 0.95 (0.85-1.07) | 0.95 (0.84-1.08) | 0.98 (0.87-1.11) | 0.88 (0.83-0.93) |

*Abbreviations:* AUD, alcohol use disorder; SVR, sustained virologic response
*Notes:* Fully adjusted model included age, sex, race/ethnicity, residence type, body mass index, smoking status, HIV co-infection, Charlson Comorbidity Index, liver-related variables (i.e., HCV genotype, hepatitis B co-infection, fibrosis 4 score, hepatic decompensation, liver cancer, and other HCV treatment-related variables (i.e., previous receipt of non-DAA therapy for HCV infection, year of DAA therapy initiation, DAA regimen type).

## Discussion

In a nationwide cohort of 94,388 middle-aged and older adults who initiated DAA therapy in the largest provider of HCV care in the United States between 2014 and 2018, we found no evidence that any level of alcohol use was associated with decreased odds of achieving SVR and this finding did not differ by baseline stage of hepatic fibrosis. Moreover, 94% of all patients achieved SVR in this “real-world” setting of an older population with multiple comorbidities who are typically underrepresented in clinical trials. Taken together, our findings support providing DAA therapy without regard to reported alcohol consumption or AUD; furthermore, payors should no longer require alcohol abstinence for reimbursement of DAA therapy for HCV infection.

Our results support the current American Association for the Study of Liver Diseases/Infectious Diseases Society of America recommendations that current or prior alcohol use is not a contraindication to HCV DAA therapy. However, a recent analysis of administrative claims and encounters in the United States found that only 23% of Medicaid, 28% of Medicare, and 35% of private insurance recipients initiated DAA treatment within one year of HCV diagnosis.^30^ Furthermore, this analysis found that Medicaid recipients were less likely to initiate DAA treatment if they resided in a state with Medicaid treatment restrictions including alcohol abstinence. Even though VA has not required alcohol abstinence prior to DAA treatment, pre-existing alcohol use still impacts who initiates DAA therapy.^31^ Given the increased risk of cirrhosis and hepatocellular carcinoma in those with chronic HCV and AUD, our findings suggest guidelines should encourage HCV treatment in those with AUD, rather than creating additional barriers to patients accessing HCV treatment. Integrating HCV treatment with AUD treatment services may prove to be a successful care model, especially because the hepatotoxic effects of alcohol on the liver have been shown to persist even after SVR.^32,33^

While numerous studies from the interferon era examined alcohol use and SVR,^1–4^ there are few such studies in the DAA era. Tsui et al examined alcohol use and SVR in the VA including the first 18 months of the DAA era.^7^ Although the authors found no difference in SVR across alcohol use categories in all primary analyses, after imputing missing HCV RNA values for 9% of their cohort, they observed a significantly lower likelihood of SVR in those reporting unhealthy alcohol use. However, Tsui et al also demonstrated that those with unhealthy alcohol use (defined as AUDIT-C ≥4) were more likely to have missing HCV RNA values and therefore assumptions required for multiple imputation may have been violated.^29^

Furthermore, Tsui et al used patients reporting abstinence as the referent group; however, this is a heterogeneous group comprising very few lifetime abstainers and the majority of whom quit drinking after alcohol-related or health problems.^8^ To avoid misclassification, especially in a cohort of middle-aged and older adults, we used lower-risk consumption as the referent group and distinguished patients reporting abstinence by whether they had been diagnosed with AUD. Results from our study after imputing missing HCV RNA values did not change conclusions from our primary analysis; these analyses were included only to enable comparisons with Tsui et al since assumptions of missing at random were not met. However, after assuming everyone with missing outcome data did not achieve SVR, persons with high-risk consumption or AUD had 12% decreased odds of achieving SVR in multivariable analysis. We know in clinical practice that not every missing SVR value is a treatment failure and in fact many persons are found to have SVR when re-engaged in care;^34^ therefore, we express caution in interpreting this single association.

This study has many strengths, including the availability of detailed, longitudinal, electronic health record data on a diverse nationwide cohort of HCV patients initiating DAA therapy and findings that were robust to multiple sensitivity analyses. Importantly, the VA has been more successful than most healthcare systems in the US in diagnosing and treating HCV infection given its unified electronic health record, nationalized healthcare system, and prioritization of HCV.^35^ The result is that the VA has far exceeded other healthcare systems^30,36–40^ by treating nearly 85% of patients with known chronic HCV infection in VA care.^10,41,42^

We also recognize possible limitations. First, owing to the observational nature of the study, a degree of uncertainty persists due to the potential for residual confounding. Nonetheless, we mitigated potential confounding by comprehensively accounting for numerous demographic and clinical characteristics as well as liver- and treatment-related variables. Second, we assessed HCV RNA measurements in the 6-month period following the end of DAA treatment to define SVR, which may have resulted in potential misclassification of some patients who may have experienced viral rebound; however, this has been shown to be an extremely rare event.^43^ Some patients may also have had their first evidence of SVR recorded beyond that window of measurement, which we would have classified as a missing outcome. Third, alcohol use measurement may have been influenced by both patient and provider-level factors,^44–47^ including under-reporting level of alcohol use due to social desirability bias,^48^ which may have resulted in misclassification of some patients with high-risk consumption at lower levels of consumption. However, the AUDIT-C tool is a widely validated questionnaire that has been part of a standardized triage practice within the VA and measured annually during routine healthcare visits since 2008. Furthermore, we categorized patients with AUD separately and used patients with lower-risk consumption as the referent category, which minimized the potential of misclassification of “sick quitters”.^8^ Fourth, we performed a complete case analysis, which will be unbiased if, conditional on model covariates, missingness is independent of the outcome.^26^ To that end, we demonstrated that missing outcome data was not strongly associated with any covariate used in the model suggesting complete case analysis was appropriate. However, we additionally performed multiple sensitivity analyses, all of which were consistent with our findings from our complete case analysis. Finally, while individuals in VA care represent a diversity of backgrounds, women represented a small proportion of individuals in the cohort. However, given the current payor restrictions requiring alcohol abstinence prior to initiating DAA therapy, studying the relationship between alcohol use and HCV treatment outcomes, including SVR, may be limited in other US healthcare settings.

In conclusion, achieving SVR has been shown to reduce the risk of post-SVR outcomes, including hepatocellular carcinoma, liver-related mortality, and all-cause mortality. Our findings suggest that DAA therapy should be provided and reimbursed despite alcohol consumption or history of alcohol use disorder. Restricting access to DAA therapy based on alcohol consumption or AUD creates an unnecessary barrier to patients accessing DAA therapy and challenges HCV elimination goals.

## Data Availability

Due to US Department of Veterans Affairs (VA) regulations and our ethics agreements, the analytic data sets used for this study are not permitted to leave the VA firewall without a data use agreement. This limitation is consistent with other studies based on VA data. However, VA data are made freely available to researchers with an approved VA study protocol. For more information, please visit https://www.virec.research.va.gov or contact the VA Information Resource Center at.

## Contributors

EC, CP, DF, VLR, and CR conceived the study. CP, DE, JT, and CR curated the data. CP and CR performed the formal analysis. DF and AJ acquired funding. EC, CP, DF, VLR, and CR designed the methodology. EC, CM, and CR managed and coordinated the project. DF, AJ, and CR procured resources to carry out the study. CP, DE, JT, and CR developed programming. VLR and CR provided oversight and leadership of the project. EC, CP, and CR prepared data visualizations. EC and CP wrote the first draft of the manuscript. All the authors wrote (reviewed and edited) the manuscript. The senior author attests that all listed authors meet authorship criteria and that no others meeting the criteria have been omitted.

## Acknowledgments

This work uses data provided by patients and collected by the VA as part of their care and support. The views and opinions expressed in this manuscript are those of the authors and do not necessarily represent those of the Department of Veterans Affairs or the United States government.

## Ethics

This study was approved by the institutional review boards of Yale University (ref #1506016006) and VA Connecticut Healthcare System (ref #AJ0013). It has been granted a waiver of informed consent and is compliant with the Health Insurance Portability and Accountability Act.

---

**eTable 1.** Covariate distribution between those with and without primary exposure data

|  | Not missing AUDIT-C<br>n=90,625 | Missing AUDIT-C<br>n=3,763 | SMD |
| --- | --- | --- | --- |
| Outcome |  |  |  |
| Sustained virologic response |  |  |  |
| No | 4,279 (4.7) | 181 (4.8) | 0.07 |
| Yes | 72,289 (79.8) | 2,900 (77.1) |  |
| Missing | 14,057 (15.5) | 682 (18.1) |  |
| Demographics |  |  |  |
| Age in years, median (IQR) |  |  |  |
| 48-54 | 5,135 (5.7) | 217 (5.8) | 0.01 |
| 55-59 | 21,826 (24.1) | 903 (24.0) |  |
| 60-64 | 34,841 (38.4) | 1,437 (38.2) |  |
| 65-73 | 28,823 (31.8) | 1,206 (32.0) |  |
| Sex |  |  |  |
| Women | 2,714 (3.0) | 102 (2.7) | 0.02 |
| Men | 87,911 (97.0) | 3,661 (97.3) |  |
| Race/ethnicity |  |  |  |
| Non-Hispanic White | 45,462 (50.2) | 2,005 (53.3) | 0.09 |
| Non-Hispanic Black | 35,960 (39.7) | 1,323 (35.2) |  |
| Hispanic | 5,002 (5.5) | 236 (6.3) |  |
| Other/missing | 4,201 (4.6) | 199 (5.3) |  |
| Residence type |  |  |  |
| Rural | 23,729 (26.2) | 930 (24.7) | 0.03 |
| Urban | 66,896 (73.8) | 2,833 (75.3) |  |
| Clinical characteristics |  |  |  |
| Body mass index |  |  |  |
| Underweight | 1,468 (1.6) | 65 (1.7) | 0.19 |
| Normal | 26,211 (28.9) | 1,165 (31.0) |  |
| Overweight | 34,284 (37.8) | 1,384 (36.8) |  |
| Obese | 27,866 (30.7) | 1,019 (27.1) |  |
| Missing | 796 (0.9) | 130 (3.5) |  |
| Smoking status |  |  |  |
| Never | 11,214 (12.4) | 481 (12.8) | 0.12 |
| Current | 60,665 (66.9) | 2,380 (63.2) |  |
| Former | 17,721 (19.6) | 804 (21.4) |  |
| Missing | 1,025 (1.1) | 98 (2.6) |  |
| HIV co-infection | 2,727 (3.0) | 198 (5.3) | 0.11 |
| Charlson Comorbidity Index |  |  |  |
| 1 | 37,310 (41.2) | 2,077 (55.2) | 0.29 |
| 2 | 23,320 (25.7) | 676 (18.0) |  |
| 3 | 8,821 (9.7) | 277 (7.4) |  |
| 4 | 8,169 (9.0) | 270 (7.2) |  |
| ≥5 | 13,005 (14.4) | 463 (12.3) |  |
| **Liver-related variables** |  |  |  |
| HCV Genotype |  |  |  |
| 1 | 74,999 (82.8) | 3,035 (80.7) | 0.11 |
| 2 | 8,417 (9.3) | 344 (9.1) |  |
| 3 | 4,974 (5.5) | 234 (6.2) |  |
| 4/5/6 | 886 (1.0) | 38 (1.0) |  |
| Missing | 1,349 (1.5) | 112 (3.0) |  |
| Hepatitis B co-infection | 1,789 (2.0) | 74 (2.0) | <0.01 |
| Fibrosis 4 score |  |  |  |
| <1.45 | 19,790 (21.8) | 734 (19.5) | 0.08 |
| 1.45-3.25 | 42,278 (46.7) | 1,719 (45.7) |  |
| >3.25 | 22,180 (24.5) | 991 (26.3) |  |
| Missing | 6,377 (7.0) | 319 (8.5) |  |
| Hepatic decompensation | 2,961 (3.3) | 135 (3.6) | 0.02 |
| Liver cancer | 1,933 (2.1) | 125 (3.3) | 0.07 |
| **Treatment-related variables** |  |  |  |
| Previous non-DAA therapy for HCV infection | 13,572 (15.0) | 585 (15.5) | 0.02 |
| Year of DAA therapy initiation |  |  |  |
| 2014 | 7,232 (8.0) | 306 (8.1) | 0.12 |
| 2015 | 25,304 (27.9) | 968 (25.7) |  |
| 2016 | 33,651 (37.1) | 1,289 (34.3) |  |
| 2017 | 18,659 (20.6) | 880 (23.4) |  |
| 2018 | 5,779 (6.4) | 320 (8.5) |  |
| DAA regimen type |  |  |  |
| Sofosbuvir/ledipasvir | 53,024 (58.5) | 2,200 (58.5) | 0.08 |
| Elbasvir/grazoprevir | 8,923 (9.8) | 317 (8.4) |  |
| Sofosbuvir/velpatasvir ± voxilaprevir | 7,286 (8.0) | 345 (9.2) |  |
| Glecaprevir/pibrentasvir | 3,609 (4.0) | 184 (4.9) |  |
| Paritaprevir/ritonavir/ombitasvir ± dasabuvir | 6,996 (7.7) | 282 (7.5) |  |
| Sofosbuvir/daclatasvir | 1,067 (1.2) | 41 (1.1) |  |
| Simeprevir/sofosbuvir | 3,107 (3.4) | 108 (2.9) |  |
| Sofosbuvir/ribavirin | 6,613 (7.3) | 286 (7.6) |  |

*Abbreviations:* AUDIT-C, Alcohol Use Disorder Identification Test - Consumption; SMD, standardized mean difference; AUD, alcohol use disorder; IQR, interquartile range; HIV, human immunodeficiency virus; HCV, hepatitis C virus; DAA, direct-acting antiviral
*Notes:* All statistics reported as n(%) unless otherwise noted.

**eTable 2.** Covariate distribution between those with and without outcome data

|  | Not missing SVR<br>n=76,568 | Missing SVR<br>n=14,057 | SMD |
| --- | --- | --- | --- |
| Exposure |  |  |  |
| Alcohol category |  |  |  |
| Abstinent with no AUD | 35,868 (46.8) | 6,055 (43.1) | 0.11 |
| Abstinent with AUD | 10,046 (13.1) | 1,958 (13.9) |  |
| Lower-risk consumption | 14,879 (19.4) | 2,632 (18.7) |  |
| Moderate-risk consumption | 3,489 (4.6) | 629 (4.5) |  |
| High-risk consumption or AUD | 12,286 (16.0) | 2,783 (19.8) |  |
| Demographics |  |  |  |
| Age in years, median (IQR) |  |  |  |
| 48-54 | 4,196 (5.5) | 939 (6.7) | 0.11 |
| 55-59 | 18,047 (23.6) | 3,779 (26.9) |  |
| 60-64 | 29,474 (38.5) | 5,367 (38.2) |  |
| 65-73 | 24,851 (32.5) | 3,972 (28.3) |  |
| Sex |  |  |  |
| Women | 2,295 (3.0) | 419 (3.0) | <0.01 |
| Men | 74,273 (97.0) | 13,638 (97.0) |  |
| Race/ethnicity |  |  |  |
| Non-Hispanic White | 38,588 (50.4) | 6,874 (48.9) | 0.12 |
| Non-Hispanic Black | 30,577 (39.9) | 5,383 (38.3) |  |
| Hispanic | 3,875 (5.1) | 1,127 (8.0) |  |
| Other/missing | 3,528 (4.6) | 673 (4.8) |  |
| Residence type |  |  |  |
| Rural | 20,070 (26.2) | 3,659 (26.0) | <0.01 |
| Urban | 56,498 (73.8) | 10,398 (74.0) |  |
| Clinical characteristics |  |  |  |
| Body mass index |  |  |  |
| Underweight | 1,178 (1.5) | 290 (2.1) | 0.12 |
| Normal | 21,761 (28.4) | 4,450 (31.7) |  |
| Overweight | 29,063 (38.0) | 5,221 (37.1) |  |
| Obese | 23,969 (31.3) | 3,897 (27.7) |  |
| Missing | 597 (0.8) | 199 (1.4) |  |
| Smoking status |  |  |  |
| Never | 9,691 (12.7) | 1,523 (10.8) | 0.15 |
| Current | 50,454 (65.9) | 10,211 (72.6) |  |
| Former | 15,569 (20.3) | 2,152 (15.3) |  |
| Missing | 854 (1.1) | 171 (1.2) |  |
| HIV co-infection | 2,354 (3.1) | 373 (2.7) | 0.03 |
| Charlson Comorbidity Index |  |  |  |
| 1 | 31,268 (40.8) | 6,042 (43.0) | 0.06 |
| 2 | 19,667 (25.7) | 3,653 (26.0) |  |
| 3 | 7,468 (9.8) | 1,353 (9.6) |  |
| 4 | 7,027 (9.2) | 1,142 (8.1) |  |
| ≥5 | 11,138 (14.5) | 1,867 (13.3) |  |
| Liver-related variables |  |  |  |
| HCV Genotype |  |  |  |
| 1 | 63,696 (83.2) | 11,303 (80.4) | 0.08 |
| 2 | 6,965 (9.1) | 1,452 (10.3) |  |
| 3 | 4,106 (5.4) | 868 (6.2) |  |
| 4/5/6 | 749 (1.0) | 137 (1.0) |  |
| Missing | 1,052 (1.4) | 297 (2.1) |  |
| Hepatitis B co-infection | 1,443 (1.9) | 346 (2.5) | 0.04 |
| Fibrosis 4 score |  |  |  |
| <1.45 | 16,566 (21.6) | 3,224 (22.9) | 0.06 |
| 1.45-3.25 | 36,068 (47.1) | 6,210 (44.2) |  |
| >3.25 | 18,656 (24.4) | 3,524 (25.1) |  |
| Missing | 5,278 (6.9) | 1,099 (7.8) |  |
| Hepatic decompensation | 2,426 (3.2) | 535 (3.8) | 0.03 |
| Liver cancer | 1,672 (2.2) | 261 (1.9) | 0.02 |
| Treatment-related variables |  |  |  |
| Previous non-DAA therapy for HCV infection | 11,649 (15.2) | 1,923 (13.7) | 0.04 |
| Year of DAA therapy initiation |  |  |  |
| 2014 | 6,305 (8.2) | 927 (6.6) | 0.19 |
| 2015 | 21,928 (28.6) | 3,376 (24.0) |  |
| 2016 | 28,560 (37.3) | 5,091 (36.2) |  |
| 2017 | 15,324 (20.0) | 3,335 (23.7) |  |
| 2018 | 4,451 (5.8) | 1,328 (9.4) |  |
| DAA regimen type |  |  |  |
| Sofosbuvir/ledipasvir | 44,945 (58.7) | 8,079 (57.5) | 0.13 |
| Elbasvir/grazoprevir | 7,682 (10.0) | 1,241 (8.8) |  |
| Sofosbuvir/velpatasvir ± voxileprevir | 5,941 (7.8) | 1,345 (9.6) |  |
| Glecaprevir/pibrentasvir | 2,843 (3.7) | 766 (5.4) |  |
| Paritaprevir/ritonavir/ombitasvir ± dasabuvir | 5,986 (7.8) | 1,010 (7.2) |  |
| Sofosbuvir/daclatasvir | 895 (1.2) | 172 (1.2) |  |
| Simeprevir/sofosbuvir | 2,748 (3.6) | 359 (2.6) |  |
| Sofosbuvir/ribavirin | 5,528 (7.2) | 1,085 (7.7) |  |

*Abbreviations:* SMD, standardized mean difference; AUD, alcohol use disorder; IQR, interquartile range; HIV, human immunodeficiency virus; HCV, hepatitis C virus; DAA, direct-acting antiviral
*Notes:* All statistics reported as n(%) unless otherwise noted.

**The RECORD statement – checklist of items, extended from the STROBE statement, that should be reported in observational studies using routinely collected health data**.

|  | Item No. | STROBE items | Location in manuscript where items are reported | RECORD items | Location in manuscript where items are reported |
| --- | --- | --- | --- | --- | --- |
| **Title and abstract** |  |  |  |  |  |
|  | 1 | (a) Indicate the study's design with a commonly used term in the title or the abstract (b) Provide in the abstract an informative and balanced summary of what was done and what was found | (a) Title & Abstract<br>(b) Abstract | RECORD 1.1: The type of data used should be specified in the title or abstract. When possible, the name of the databases used should be included.<br>RECORD 1.2: If applicable, the geographic region and timeframe within which the study took place should be reported in the title or abstract.<br>RECORD 1.3: If linkage between databases was conducted for the study, this should be clearly stated in the title or abstract. | 1.1: Abstract – Design<br>1.2: Abstract – Setting & Patients<br>1.3: n/a |
| **Introduction** |  |  |  |  |  |
| Background rationale | 2 | Explain the scientific background and rationale for the investigation being reported | Background |  |  |
| Objectives | 3 | State specific objectives, including any prespecified hypotheses | Background (End of Para 3) |  |  |
| **Methods** |  |  |  |  |  |
| Study Design | 4 | Present key elements of study design early in the paper | Methods (Study design and data source & Study population) |  |  |
| Setting | 5 | Describe the setting, locations, and relevant dates, including | Methods (Study design and data |  |  |
|  |  | periods of recruitment, exposure, follow-up, and data collection | source; Study population; Alcohol category; Outcome and follow-up) |  |  |
| Participants | 6 | (a) *Cohort study* - Give the eligibility criteria, and the sources and methods of selection of participants. Describe methods of follow-up<br>*Case-control study* - Give the eligibility criteria, and the sources and methods of case ascertainment and control selection. Give the rationale for the choice of cases and controls<br>*Cross-sectional study* - Give the eligibility criteria, and the sources and methods of selection of participants<br>(b) *Cohort study* - For matched studies, give matching criteria and number of exposed and unexposed<br>*Case-control study* - For matched studies, give matching criteria and the number of controls per case | (a) Methods (Study population)<br>(b) n/a | RECORD 6.1: The methods of study population selection (such as codes or algorithms used to identify subjects) should be listed in detail. If this is not possible, an explanation should be provided.<br>RECORD 6.2: Any validation studies of the codes or algorithms used to select the population should be referenced. If validation was conducted for this study and not published elsewhere, detailed methods and results should be provided.<br>RECORD 6.3: If the study involved linkage of databases, consider use of a flow diagram or other graphical display to demonstrate the data linkage process, including the number of individuals with linked data at each stage. | 6.1: Methods (Study population)<br>6.2: Methods (Study population; Alcohol category; Outcome; Covariates)<br>6.3: n/a |
| Variables | 7 | Clearly define all outcomes, exposures, predictors, potential confounders, and effect modifiers. Give diagnostic criteria, if applicable. | Methods (Alcohol category; Outcome; Covariates; Statistical analyses) | RECORD 7.1: A complete list of codes and algorithms used to classify exposures, outcomes, confounders, and effect modifiers should be provided. If these cannot be reported, an explanation should be provided. | 7.1: Methods (Covariates) |
| Data sources/<br>measurement | 8 | For each variable of interest, give sources of data and details of methods of assessment (measurement). Describe comparability of assessment methods if there is more than one group | Methods (Alcohol category; Outcome; Covariates) |  |  |
| Bias | 9 | Describe any efforts to address potential sources of bias | Methods (Statistical analysis; Sensitivity analyses) |  |  |
| Study size | 10 | Explain how the study size was arrived at | Methods (Study design and data source; Study population; Statistical analysis) |  |  |
| Quantitative variables | 11 | Explain how quantitative variables were handled in the analyses. If applicable, describe which groupings were chosen, and why | Methods (Statistical analysis) |  |  |
| Statistical methods | 12 | (a) Describe all statistical methods, including those used to control for confounding<br>(b) Describe any methods used to examine subgroups and interactions<br>(c) Explain how missing data were addressed<br>(d) *Cohort study* - If applicable, explain how loss to follow-up was addressed<br>*Case-control study* - If applicable, explain how matching of cases and controls was addressed | (a-c) Methods (Covariates; Statistical analysis; Sensitivity analyses)<br>(d) n/a<br>(e) Methods (Sensitivity analyses) |  |  |
|  |  | *Cross-sectional study* - If applicable, describe analytical methods taking account of sampling strategy<br>(e) Describe any sensitivity analyses |  |  |  |
| Data access and cleaning methods |  | .. |  | RECORD 12.1: Authors should describe the extent to which the investigators had access to the database population used to create the study population.<br>RECORD 12.2: Authors should provide information on the data cleaning methods used in the study. | 12.1: Methods (Role of the funding source)<br>12.2: Methods (Alcohol category; Outcome; Covariates; Statistical analysis) |
| Linkage |  | .. |  | RECORD 12.3: State whether the study included person-level, institutional-level, or other data linkage across two or more databases. The methods of linkage and methods of linkage quality evaluation should be provided. | n/a |
| **Results** |  |  |  |  |  |
| Participants | 13 | (a) Report the numbers of individuals at each stage of the study (*e.g.*, numbers potentially eligible, examined for eligibility, confirmed eligible, included in the study, completing follow-up, and analysed)<br>(b) Give reasons for non-participation at each stage.<br>(c) Consider use of a flow diagram | (a-c) Results (Cohort description) | RECORD 13.1: Describe in detail the selection of the persons included in the study (*i.e.*, study population selection) including filtering based on data quality, data availability and linkage. The selection of included persons can be described in the text and/or by means of the study flow diagram. | 13.1: Results (Cohort description) |
| Descriptive data | 14 | (a) Give characteristics of study participants (*e.g.*, demographic, clinical, social) and information on exposures and potential confounders<br>(b) Indicate the number of participants with missing data for each variable of interest<br>(c) *Cohort study* - summarise follow-up time (*e.g.*, average and total amount) | (a) Results (Cohort description, Table 1, eTables 1 & 2)<br>(b) Table 1, eTables 1 & 2<br>(c) Results (Cohort description) |  |  |
| Outcome data | 15 | *Cohort study* - Report numbers of outcome events or summary measures over time<br>*Case-control study* - Report numbers in each exposure category, or summary measures of exposure<br>*Cross-sectional study* - Report numbers of outcome events or summary measures | Results (Alcohol category and SVR) |  |  |
| Main results | 16 | (a) Give unadjusted estimates and, if applicable, confounder-adjusted estimates and their precision (*e.g.*, 95% confidence interval). Make clear which confounders were adjusted for and why they were included<br>(b) Report category boundaries when continuous variables were categorized<br>(c) If relevant, consider translating estimates of relative risk into absolute risk for a meaningful time period | (a) Results (Alcohol category and SVR; Table 2)<br>(b) Results (Tables 1, 2, and 3)<br>(c) Results (Alcohol category and SVR) |  |  |
| Other analyses | 17 | Report other analyses done—<br>e.g., analyses of subgroups and interactions, and sensitivity analyses | Results (Alcohol category and SVR; Sensitivity analyses) |  |  |
| **Discussion** |  |  |  |  |  |
| Key results | 18 | Summarise key results with reference to study objectives | Discussion (Para 1) |  |  |
| Limitations | 19 | Discuss limitations of the study, taking into account sources of potential bias or imprecision. Discuss both direction and magnitude of any potential bias | Discussion (Para 5 and 6) | RECORD 19.1: Discuss the implications of using data that were not created or collected to answer the specific research question(s). Include discussion of misclassification bias, unmeasured confounding, missing data, and changing eligibility over time, as they pertain to the study being reported. | 19.1 Discussion (Para 5 and 6) |
| Interpretation | 20 | Give a cautious overall interpretation of results considering objectives, limitations, multiplicity of analyses, results from similar studies, and other relevant evidence | Discussion (Throughout) |  |  |
| Generalisability | 21 | Discuss the generalisability (external validity) of the study results | Discussion (Para 5 and 6) |  |  |
| **Other Information** |  |  |  |  |  |
| Funding | 22 | Give the source of funding and the role of the funders for the present study and, if applicable, for the original study on which the present article is based | Methods (Role of the funding source) |  |  |
| Accessibility of protocol, raw data, and |  | .. |  | RECORD 22.1: Authors should provide information on how to access any supplemental information such as | Data availability statement |
| programming<br>code |  |  |  | the study protocol, raw data, or<br>programming code. |  |

\*Reference: Benchimol EI, Smeeth L, Guttman A, Harron K, Moher D, Petersen I, Sørensen HT, von Elm E, Langan SM, the RECORD Working Committee. The REporting of studies Conducted using Observational Routinely-collected health Data (RECORD) Statement. *PLoS Medicine* 2015; in press.
\*Checklist is protected under Creative Commons Attribution ([CC BY](https://creativecommons.org/licenses/by/4.0/)) license.

